# Characterizing Loneliness and Health in US Adults: An analysis of 2024 National Health Interview Survey

**DOI:** 10.64898/2026.04.16.26351034

**Authors:** Troy C. Dildine, Colleen Burke, Flavia P. Kapos

**Author notes:** Corresponding Author: Troy C. Dildine, Ste 200, 1070 Arastradero Rd., Palo Alto, CA, 650-723- 3032. Data availability: National Health Interview Survey data are made public at the following: https://www.cdc.gov/nchs/nhis/documentation/2024-nhis.html.

## Abstract

**Background:** Loneliness is common and deleterious to health. Yet little is known about its population burden and health correlates in the US. We aimed to determine the prevalence of loneliness and characterize its health and social functioning correlates among US adults.

**Methods:** With data from the National Health Interview Study (2024), we used survey-weighted Poisson regression to estimate relative risks (RR) and 95% confidence intervals (CI) for frequent loneliness by levels of self-reported general health, social/emotional support, social functioning, and healthcare utilization, adjusted for age, sex, race/ethnicity, number of people in household, marital status, and psychological distress.

**Results:** 12 million US adults reported usually or always feeling lonely, which was associated with worse general health and social/emotional support, work and social participation limitations, and healthcare disengagement.

**Conclusions:** Loneliness affects millions of US adults, with substantial health and social functioning burden.

## Introduction

Federal campaigns in the United States^1^ and the UK^2^ highlight the attention loneliness has garnered; yet, only the UK has assessed loneliness’ national burden^3-4^, with little known quantitatively about loneliness’ impact and health burden in the US. This reflects a broader trend of underrepresentation of social measures in medicine despite growing evidence loneliness has widespread impacts in health in both^5^ human^6-7^ and animal^8-9^ models.

Social determinants of health have provided a framework for understanding how individual, interpersonal, group, and societal level factors can impact health^10-13^. Among these factors, loneliness is one of the most deleterious, with greater mortality risk than obesity and frequent daily cigarette use^14^. Loneliness is a subjective feeling of being alone (i.e., perceived isolation) or not feeling socially connected with others^15^ that is separate from but can include objective isolation. In 2024, the National Health Interview Survey (NHIS^16^) included a measure of loneliness for the first time in its annual report. Capitalizing on this new nationally representative data to enhance external validity^17^, we sought to 1) determine the prevalence of loneliness among US adults, and 2) characterize the burden of loneliness on indicators of health and social functioning.

We hypothesized that individuals who reported feeling lonely would report worse social functioning, emotional and social support, and general health compared to those who report less or no experiences of loneliness. Secondarily, we hypothesized that lonely individuals would exhibit a bimodal distribution in their healthcare utilization patterns (i.e., some would seek healthcare for social connection whereas others without proper support would remain isolated).

## Methods

### Sample

We analyzed data from the 2024 NHIS, a nationally representative survey of US non-institutionalized civilian adults collected via in-home and telephone interviews.

### Questionnaires

<u>Loneliness</u> was assessed using a five-item Likert scale with the following question, “How often do you feel lonely?” with the following response options, “always”, “usually”, “sometimes”, “rarely”, and “never.” We operationalized frequent loneliness as anyone who endorsed feeling lonely “usually” or “always” and compared this with individuals who endorsed feeling lonely, “sometimes”, “rarely” or “never” (see Supplement eMethods for more details).

In addition to our outcome of interest, we estimated how loneliness was associated with several health and social functioning measures.

#### Emotional and social support

“How often do you get the social and emotional support you need?” (“always”, “usually”, “sometimes”, “rarely”, and “never.”)

#### Social functioning

“Because of a physical, mental, or emotional condition, do you have difficulty doing errands alone such as visiting a doctor’s office or shopping?”

“Because of a physical, mental, or emotional condition, do you have difficulty participating in social activities such as visiting friends, attending clubs and meetings, or going to parties?” “Are you limited in the kind OR amount of work you can do because of a physical, mental or emotional problem?”

#### Self-reported general health

“Would you say your health in general is excellent, very good, good, fair, or poor?” Arthritis and rheumatic disorders:

“Has a health professional diagnosed you with some form of arthritis, rheumatoid arthritis, gout, lupus, or fibromyalgia?” (‘yes’, ‘no’, ‘refused’, and ‘don’t know’).

#### Healthcare utilization

“About how long has it been since you last saw a doctor or other health professional about your health?” (“never”, “past year”, “last 2 years”, “3 years”, “5 years”, “10 years”, “more than 10 years”, “refused”, and “don’t know”).

### Data analysis

We preregistered our analysis at AsPredicted: (link_available_on_publication). Survey weighting was applied to all analyses using the survey package in Stata. Multivariable Poisson regression was used to estimate the associations between loneliness and our dependent variables, accounting for a priori covariates. We computed survey-weighted relative risk scores with 95% confidence intervals. Adjustment covariates included psychological distress, age (in years, top-coded at 85), sex, race/ethnicity, number of persons in household (0-6+), and marital status (more details in Table 1). Sensitivity analyses included an alternative loneliness definition by including the “sometimes” responders to the lonely group. A total of 579 individuals were excluded from our analyses due to missing data in study variables (1.8% unweighted, 1.7% weighted).

**Table 1.** Survey-weighted population estimates of characteristics by presence of loneliness.

|  | **Loneliness<sup>a</sup>** |  |
| --- | --- | --- |
|  | **No** | **Yes** |
| Unweighted sample size | n=29,736 | n=1,734 |
| **Population estimate** | 238 million | 12 million |
|  | (95.1%) | (4.9%) |
| **Age in years, mean (SD)** | 48.4 (18.6) | 45.9 (18.9) |
| **Race/Ethnicity, %** |  |  |
| Hispanic | 17.9 | 18.1 |
| Non-Hispanic (NH) American Indian or Alaska Native | 1.2 | 3.3 |
| NH Asian only | 6.3 | 4.9 |
| NH Black/AA only | 11.4 | 13.6 |
| NH White only | 61.8 | 58.0 |
| NH other single or multiple races | 1.5 | 2.2 |
| **Sex, %** |  |  |
| Female | 51.3 | 53.2 |
| Male | 48.7 | 46.8 |
| **Marital Status, %** |  |  |
| Married or Living with Partner | 62.4 | 34.7 |
| Single/Never Married | 23.1 | 38.8 |
| Widowed, Separated, Divorced | 14.5 | 26.6 |
| **Psychological distress (K-6 scale, 0-24), mean (SD)** | 2.5 (3.4) | 8.8 (6.4) |
| **Serious psychological distress (K-6), %** |  |  |
| Yes ($\geq 13$ points) | 2.4 | 30.4 |
| No ($< 13$ points) | 97.6 | 69.6 |
| **# Persons in Household, mean (SD)** | 2.7(1.2) | 2.5 (1.3) |

<sup>a</sup> Adults who reported feeling lonely “always” or “usually” were considered to experience loneliness (vs. “sometimes, “rarely”, or “never”).

## Results

### Prevalence and sample characteristics of loneliness in the US

Among 31,470 adults representing 250 million US adults, approximately 2.5% (6.2 million) reported feeling lonely always, 2.4% (6 million) feel lonely usually, 18.8% (47.1 million) feel lonely sometimes, 25.7% (64.3 million) rarely feel lonely, and 50.6% (126.4 million) never feel lonely (see Figure 1). For our main analysis, with a more conservative definition of loneliness (usually or always), 12 million US adults met this operationalization. Lonely adults were slightly younger (mean 45.9 vs. 48.4 years) and less likely to be married/partnered (34.7% vs. 62.4%). Psychological distress was markedly elevated among lonely adults: mean K-6 score 8.8 (SD=6.4) versus 2.5 (SD=3.4), with 30.4% meeting serious psychological distress criteria versus 2.4% among non-lonely adults. Race and sex distributions were generally similar between groups (full results in Table 1).

**Figure 1.**
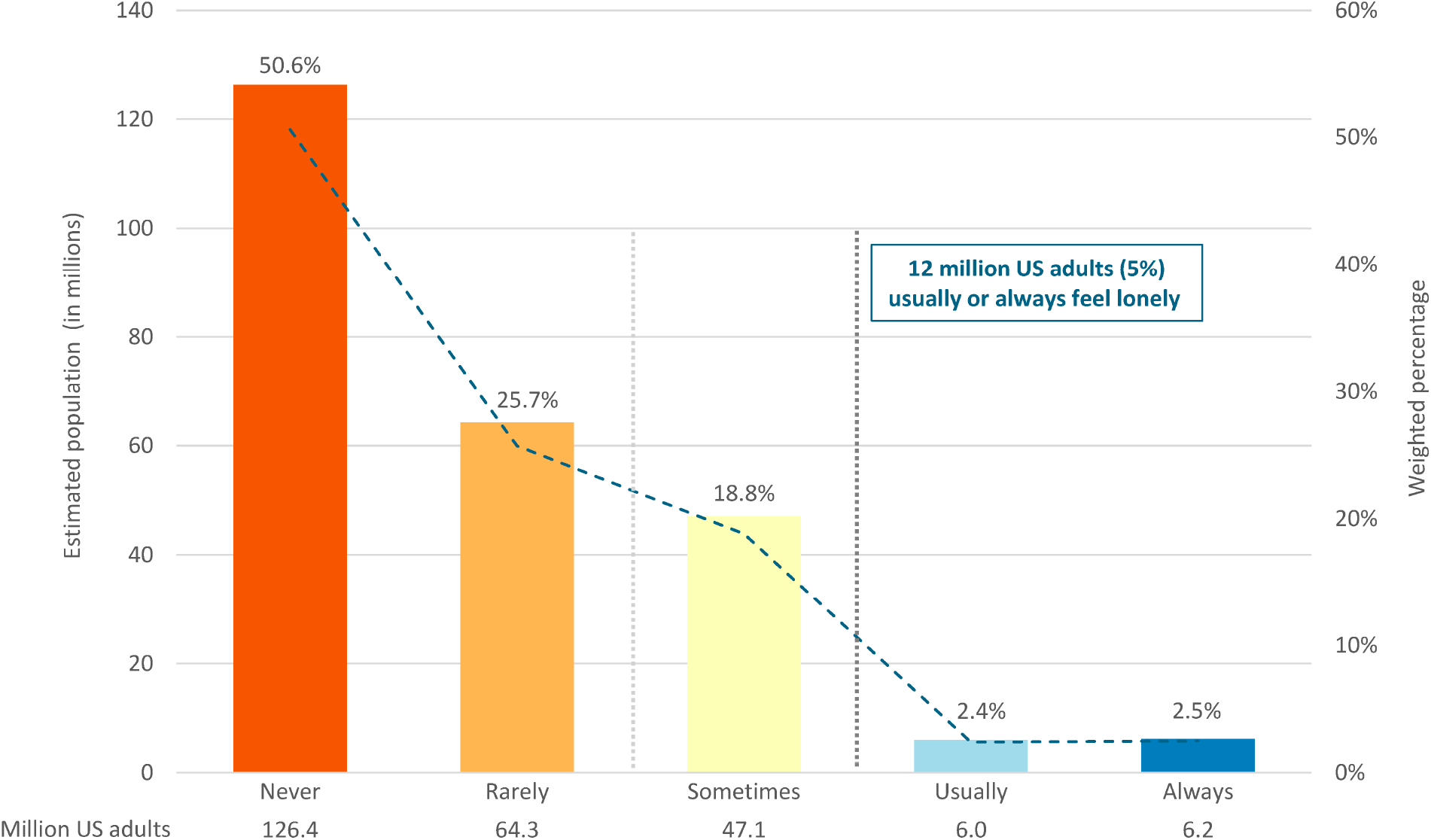
Prevalence of Loneliness in US Adults, NHIS 2024 1. Bar graphs demonstrate the weighted estimates for how many US noninstitutionalized civilian adults endorsed each of the response categories to the question, ‘How often do you feel lonely?’ Going from right (more loneliness) to left, approximately 2.5% (6.2 million) reported feeling lonely always, 2.4% (6 million) feel lonely usually, 18.8% (47.1 million) feel lonely sometimes, 25.7% (64.3 million) rarely feel lonely, and 50.6% (126.4 million) never feel lonely. On the y-axis on the left we show estimated population in millions and report this underneath each bar and on the y-axis on the right of the grid, we report weighted percentages (i.e., up to 100% of US noninstitutionalized civilians adults) with percentages above each bar.

### Loneliness is associated with less social support, greater limitation in work and social participation, and worse general health

After adjusting for age, sex, race/ethnicity, number of people in household, marital status, and psychological distress, several factors were significantly associated with loneliness. Social/emotional support had the strongest association (p<0.001): adults who “rarely” had support (RR=4.52, 95%CI: 3.82, 5.36), and those who “never” had support (RR=3.12, 95%CI: 2.58, 3.77) were substantially more likely to be lonely (full results in Table 2). Social participation difficulty was significantly associated with loneliness (p<0.001): adults with “some difficulty” (RR=1.43, 95% CI: 1.23, 1.67) and “a lot of difficulty” (RR=1.26, 95%CI: 1.05, 1.52) were more likely to be lonely. Adults who had work limitations (RR=1.21, 95% CI: 1.05, 1.39; p=0.010) were also more likely to be lonely, while there was no evidence of association between difficulty running errands alone and loneliness (p=0.381). We also demonstrated a dose-effect association between self-reported general health status and loneliness (e.g., fair health RR=1.54, 95%CI: 1.21, 1.98; poor health RR=1.44, 95%CI: 1.08, 1.91; vs. excellent health, p=0.001). Further, time since last seeing a health professional showed a significant association (p=0.027), with adults not seen in 10+ years more likely to be lonely (RR=1.79, 95%CI: 1.15, 2.77, vs. those seen within the past 12 months).

**Table 2.** Multivariable Poisson regression estimates of associations with loneliness.

| **Loneliness<sup>a</sup>** |  |  |  |
| --- | --- | --- | --- |
| Unweighted n= 30,891 |  |  |  |
| Weighted pop.= 246 million |  |  |  |
|  | **RR** | **95% CI** | **p-value** |
| **Social and emotional support** |  |  | **<0.001** |
| Always | Reference | Reference |  |
| Usually | 1.19 | 0.99, 1.44 |  |
| Sometimes | 2.38 | 1.99, 2.85 |  |
| Rarely | 4.52 | 3.82, 5.36 |  |
| Never | 3.12 | 2.58, 3.77 |  |
| **Social functioning: Errands Alone** |  |  | **0.381** |
| No difficulty | Reference | Reference |  |
| Some difficulty | 1.13 | 0.96, 1.33 |  |
| A lot of difficulty | 0.93 | 0.74, 1.18 |  |
| Cannot do at all | 1.01 | 0.76, 1.98 |  |
| **Social functioning: Social Participation** |  |  | **<0.001** |
| No difficulty | Reference | Reference |  |
| Some difficulty | 1.43 | 1.23, 1.67 |  |
| A lot of difficulty | 1.26 | 1.05, 1.52 |  |
| Cannot do at all | 1.11 | 0.83, 1.49 |  |
| **Social functioning: Work Limitations** |  |  | **0.010** |
| No | Reference | Reference |  |
| Yes | 1.21 | 1.05, 1.39 |  |
| **General health** |  |  | **0.001** |
| Excellent | Reference | Reference |  |
| Very good | 1.12 | 0.88, 1.41 |  |
| Good | 1.42 | 1.13, 1.77 |  |
| Fair | 1.54 | 1.21, 1.98 |  |
| Poor | 1.44 | 1.08, 1.91 |  |
| **Diagnosis of arthritis** |  |  | **0.195** |
| No | Reference | Reference |  |
| Yes | 1.10 | 0.95, 1.26 |  |
| **Time since last saw health professional** |  |  | **0.027** |
| Within the past year | Reference | Reference |  |
| Within the past 2 years | 0.96 | 0.77, 1.21 |  |
| Within the past 3 years | 1.05 | 0.73, 1.49 |  |
| Within the past 5 years | 1.01 | 0.70, 1.45 |  |
| Within the past 10 years | 1.22 | 0.75, 1.99 |  |
| 10 or more years | 1.79 | 1.15, 2.77 |  |
| Never | 0.08 | 0.01, 0.57 |  |
| **Health visit within the past 12 months** |  |  | **0.546** |
| Yes | Reference | Reference |  |
| No | 1.05 | 0.89, 1.24 |  |

<sup>a</sup>Adults who reported feeling lonely “always” or “usually” were considered to experience loneliness (vs. “sometimes”, “rarely”, or “never”). RR= relative risk; CI=Confidence Interval; p-values obtained from joint Wald tests. All estimates are adjusted for age, sex, race/ethnicity, number of persons in household, marital status, and psychological distress. Bolded p-values indicate statistical significance at $\alpha=0.05$ .

#### Sensitivity Analysis

Broadening loneliness’ definition to include “sometimes” increased prevalence to 23.7% (59 million adults). This revealed similar demographic patterns (see Supplement eTable 1) and multivariable associations remained consistent (see Supplement eTable 2).

## Discussion

Using the first nationally representative US data on loneliness, we found that approximately 12 million American adults (4.9%) experience frequent loneliness (“usually” or “always”), with an additional 47 million (18.8%) experiencing occasional loneliness (“sometimes”). These estimates establish the substantial public health burden of loneliness in the US, validating its recent attention^1^.

Our multivariable analyses, adjusting for psychological distress and sociodemographic factors, revealed several important patterns. The relationship between lower social/emotional support and loneliness (RRs up to 4.52) supports that these are strongly related but distinct constructs, as loneliness reflects subjective experiences while support availability may represent perceived aid and resources received. Even those “sometimes” lacking social/emotional support had elevated loneliness risk, suggesting that even unstable/unreliable support may be problematic. We also observed dose-response associations between worse general health and loneliness even in the fully adjusted models (i.e., controlling for psychological distress), suggesting that health correlates of loneliness go beyond mental health symptoms.

However, the cross-sectional design precludes determining whether poor health leads to loneliness, or whether loneliness contributes to health deterioration through a host of potential mechanisms (e.g., healthcare underutilization). Contrary to our hypotheses, there was no evidence of some lonely adults engaging in higher healthcare utilization. The finding that adults not seeing health professionals for 10+ years showed 79% higher loneliness risk has multiple potential interpretations. Loneliness may create barriers to care-seeking through lack of social support to facilitate and attend appointments. This pattern warrants longitudinal investigation to determine directionality and inform future interventions.

Our findings suggest multiple possible intervention targets for loneliness. For example, healthcare systems could screen for loneliness, particularly among unmarried individuals, those with functional limitations, and infrequent healthcare users. The large overlap with psychological distress (30.4% vs. 2.4% serious distress prevalence) suggests mental health services should routinely assess loneliness, while loneliness interventions should incorporate mental health support – in line with previous literature^18-19^.

### Limitations

The cross-sectional design hampers direct causal inference and the single-item loneliness measure, while brief and feasible for population surveillance, lacks the depth of multidimensional loneliness scales like the UCLA Loneliness Scale. Our 4.9% prevalence using stringent criteria (“usually/always”) is lower than some studies using broader definitions, though our sensitivity analysis (23.7% including “sometimes”) aligns more with UCLA-Loneliness cutoffs for loneliness (scores averaging below ‘sometimes’).

## Conclusion

Loneliness affects millions of Americans and associates with worse social functioning, health, and healthcare engagement independent of psychological distress and sociodemographics. These findings establish baseline prevalence and correlates, supporting public health response and informed intervention development.

## Data Availability

National Health Interview Survey data are made public at the following: https://www.cdc.gov/nchs/nhis/documentation/2024-nhis.html

