## Supplement for "Characterizing Loneliness and Health in US Adults: An analysis of 2024 National Health Interview Survey"

eMethods.

As noted in the main manuscript, loneliness was assessed in the NHIS using the following question, “How often do you feel lonely?” with the following response options, “always”, “usually”, “sometimes”, “rarely”, and “never.”

We attempted to be conservative with our loneliness operationalization, similar to other fields of study identifying ‘high-impact’ patients^1^. Therefore, we categorized anyone who endorsed feeling lonely “usually” or “always” as lonely. However, we note that other measures of loneliness, like the UCLA loneliness scale^2^ classify individuals as lonely using cutoffs between their rarely and sometimes range (39-53^3^) on a 20-80 scale range. A score of 60 would be associated with choosing ‘sometimes’ for every option, indicating that a response of ‘sometimes’ would easily be considered as lonely in the UCLA scale context. Given this, we thought it prudent to also include sensitivity analyses (eTable 1, 2) that included individuals who reported feeling lonely ‘sometimes’ in our lonely category.

eTable 1. Survey-weighted population estimates of characteristics by the presence of loneliness (alternative definition).

|  | **Loneliness (alternative)^a^** | |
| --- | --- | --- |
|  | **No** | **Yes** |
| Unweighted sample size | n=23,096 | n=8,374 |
| **Population estimate** | 191 million  (76.3%) | 59 million  (23.7%) |
| **Age in years, mean (SD)** | 48.7 (18.4) | 47.0 (19.4) |
| **Race/Ethnicity, %** |  |  |
| Hispanic | 18.4 | 16.3 |
| Non-Hispanic (NH) American Indian or Alaska Native | 1.2 | 1.8 |
| NH Asian only | 6.2 | 6.3 |
| NH Black/African American only | 11.1 | 13.0 |
| NH White only | 61.9 | 60.6 |
| NH other single or multiple races | 1.4 | 1.9 |
| **Sex, %** |  |  |
| Female | 49.6 | 57.3 |
| Male | 50.4 | 42.7 |
| **Marital Status, %** |  |  |
| Married or Living with Partner | 66.7 | 42.6 |
| Single/Never Married | 20.8 | 33.6 |
| Widowed, Separated, Divorced | 12.4 | 23.8 |
| **Psychological distress (K-6 scale, 0-24), mean (SD)** | 1.8 (2.8) | 5.8 (5.2) |
| **Serious psychological distress (K-6), %** |  |  |
| Yes (≥13 points) | 1.1 | 12.6 |
| No (<13 points) | 98.9 | 87.4 |
| **# Persons in Household, mean (SD)** | 2.8 (1.2) | 2.5(1.2) |

^a^ Adults who reported feeling lonely “always”, “usually”, or “sometimes” were considered to experience loneliness (vs. “rarely” or “never”).

eTable 2. Multivariable Poisson regression estimates of associations with loneliness (alternative definition).

| **Loneliness^a^** |  |  |  |
| --- | --- | --- | --- |
| Unweighted n= 30,891  Weighted pop.= 246 million | **RR** | **95% CI** | **p-value** |
| **Social and emotional support** |  |  | **<0.001** |
| Always | Reference | Reference |  |
| Usually | 1.84 | 1.73, 1.96 |  |
| Sometimes | 2.73 | 2.56, 2.90 |  |
| Rarely | 2.47 | 2.29, 2.67 |  |
| Never | 1.64 | 1.48, 1.83 |  |
| **Social functioning: Errands Alone** |  |  | **<0.001** |
| No difficulty | Reference | Reference |  |
| Some difficulty | 1.09 | 1.01, 1.17 |  |
| A lot of difficulty | 0.78 | 0.69, 0.89 |  |
| Cannot do at all | 1.01 | 0.90, 1.14 |  |
| **Social functioning: Social Participation** |  |  | **<0.001** |
| No difficulty | Reference | Reference |  |
| Some difficulty | 1.30 | 1.22, 1.38 |  |
| A lot of difficulty | 0.94 | 0.85, 1.03 |  |
| Cannot do at all | 0.98 | 0.85, 1.13 |  |
| **Social functioning: Work Limitations** |  |  | **<0.001** |
| No | Reference | Reference |  |
| Yes | 1.14 | 1.08, 1.21 |  |
| **General health** |  |  | **<0.001** |
| Excellent | Reference | Reference |  |
| Very good | 1.31 | 1.20, 1.43 |  |
| Good | 1.53 | 1.41, 1.65 |  |
| Fair | 1.54 | 1.41, 1.69 |  |
| Poor | 1.41 | 1.24, 1.60 |  |
| **Diagnosis of arthritis** |  |  | 0.116 |
| No | Reference | Reference |  |
| Yes | 1.05 | 0.99, 1.11 |  |
| **Time since last saw health professional** |  |  | 0.345 |
| Within the past year | Reference | Reference |  |
| Within the past 2 years | 1.02 | 0.94, 1.12 |  |
| Within the past 3 years | 1.20 | 1.03, 1.39 |  |
| Within the past 5 years | 1.00 | 0.85, 1.19 |  |
| Within the past 10 years | 1.09 | 0.89, 1.33 |  |
| 10 or more years | 1.08 | 0.57, 2.05 |  |
| Never |  |  | 0.068 |
| **Health visit within the past 12 months** | Reference | Reference |  |
| Yes | 1.06 | 1.00, 1.14 |  |
| No |  |  |  |

^a^Adults who reported feeling lonely “always”, “usually”, or “sometimes” were considered to experience loneliness (vs. “rarely”, or “never”). RR= relative risk; CI=Confidence Interval; p-values obtained from joint Wald tests. All estimates are adjusted for age, sex, race/ethnicity, number of persons in household, marital status, and psychological distress. Bolded p-values indicate statistical significance at α=0.05.
